# Sleep quality, mental health and circadian rhythms during COVID lockdown – Results from the SleepQuest Study

**DOI:** 10.1101/2020.07.08.20148171

**Authors:** Neil Carrigan, Alfie Wearn, Saba Meky, James Selwood, Hugh D. Piggins, Nicholas Turner, Rosemary Greenwood, Elizabeth Coulthard, On behalf of RESTED study group

## Abstract

Behavioural responses to COVID-19 lockdown will define the long-term impact of psychological stressors on sleep and brain health. Here we tease apart factors that help protect against sleep disturbance. We capitalise on the unique restrictions during COVID-19 to understand how time of day of daylight exposure and outside exercise interact with chronotype and sleep quality. 3474 people from the UK (median age 62, range 18-91) completed our online ‘SleepQuest’ Study between 29th April and 13th May 2020 – a set of validated questionnaires probing sleep quality, depression, anxiety and attitudes to sleep alongside bespoke questions on the effect of COVID-19 lockdown on sleep, time spent outside and exercising and self-help sleep measures. Significantly more people (n=1252) reported worsened than improved sleep (n=562) during lockdown (p<0.0001). Factors significantly associated with worsened sleep included low mood (p<0.001), anxiety (p<0.001) and suspected, proven or at risk of COVID-19 symptoms (all p<0.03). Sleep improvement was related to the increased length of time spent outside (P<0.01). Older people’s sleep quality was less affected than younger people by COVID lockdown (p<0.001). Better sleep quality was associated with going outside and exercising earlier, rather than later, in the day. However, the benefit of being outside early is driven by improved sleep in ‘owl’ (p=0.0002) and not ‘lark’ (p=0.27) chronotype, whereas, the benefit of early exercise (inside or outside) did not depend on chronotype. Defining the interaction between chronotype, mental health and behaviour will be critical for targeted lifestyle adaptations to protect brain health through current and future crises.

## Introduction

Poor sleep is associated with negative health outcomes including infection risk ^1^, cardiovascular disease ^2^, mental illness ^3^ and dementia ^4^. The ‘lockdown’ of normal daily life due to the COVID-19 pandemic is a (hopefully) unique circumstance in our lifetimes when daylight exposure and physical activity are constrained and much of the population is experiencing stress. Anxiety and depression play a major role in sleeplessness ^5,6^ and are linked to unhelpful attitudes and beliefs about sleep (such as “I am worried I may lose control over my abilities to sleep”) ^7,8^. Evidence so far suggests anxiety about the COVID-19 virus and its impact on society is having a detrimental effect on sleep and circadian rhythms ^9-11^.

Sleeplessness can persist even when the initial stressor has resolved ^12^. So, while COVID-19 may be a current cause of large-scale sleep disruption in the general population due to stress and anxiety, its impact on sleep and general health, may be felt long after the virus itself has largely left our communities. As we do not know how long we will be in and out of lockdown due to COVID-19, understanding the relationships between lockdown conditions, lifestyle, mental health and sleep are critical to preserve long-term mental health.

Physical activity ^13,14^ and effect of bright daylight on circadian rhythms ^15^ are known to regulate sleep and both are likely to be restricted during lockdown. UK government rules from March-May 2020 that no one should leave the house for exercise more than once a day offer a uniquely regulated period allowing interrogation of relationship between duration and timing of going outside to exercise and sleep quality. Many individuals express a behavioural preference for the waking early (‘larks’) or going to sleep late (‘owls’) as a result of interaction between endogenous circadian fluctuations in hormones and environmental factors including light ^16^. Furthermore, ‘owl’ or ‘lark’ chronotype might bias the effect of lockdown restrictions on sleep^17^. Understanding such relationships will help optimise future behavioural interventions to enhance sleep.

People over 70 may well be affected by lockdown for the longest period due to being seen as an “at risk” group and the strong suggestion that they self-isolate during lockdown. Fragmented sleep is common in later life ^18^ due to aging and the prevalence of medical problems such as sleep apnoea and chronic pain and this is associated with impaired brain health. There is a multidimensional relationship between sleep disturbance and Alzheimer’s Disease (AD) progression – not only does poor sleep increase the risk of AD ^4,19^, but AD worsens sleep ^20^ and poor sleep augments cognitive impairment found early in AD. Furthermore, when people with AD do not sleep well, they can disturb partners, who are often carers. Insomnia is a cause of carer burden and can trigger the need for institutional care ^20^. Therefore, we targeted this study to older people including those with dementia, but allowed anyone to take part as data from other groups will help us learn whether lockdown differentially affects people according to age.

Although lockdown stress might worsen sleep, we hypothesise that people might also be particularly receptive to advice, particularly if it is crowdsourced. Therefore, we felt it important to find out how people were managing sleep successfully so that we could share the knowledge with others who might be struggling.

We used an online questionnaire to test specific hypotheses:

1. COVID-related stress, anxiety, attitudes towards sleep and depression will combine to worsen average sleep quality during lockdown;
2. Sleep is generally worse in people with Mild Cognitive Impairment (MCI), dementia and people with caring responsibilities than an age-matched control population;
3. Restricted outside physical activity during lockdown will allow exploration of the interaction between time of day of outside exercise and sleep.

We also explored how people had helped their own sleep during lockdown and planned to share ideas online to enable people to take advantage of successful strategies employed by others.

## Method

### Design

The study was a cross-sectional web-based survey hosted on the REDCap system at the University of Bristol, UK. It was open to all people within the UK over the age of 18yrs between 29^th^ April and 13^th^ May 2020 (the day lockdown rules were relaxed in the UK to allow more than one trip outside). After arriving at the study home page, participants read information detailing the aims of the study, the type of data being collected and how it would be stored, anonymously analysed and published. They were then asked to tick a box to consent to taking part before continuing to the questionnaire. It took on average 20mins to complete the survey.

Recruitment to the study was primarily through the Join Dementia Research (JDR) database which sent out an email message informing participants of the study with a link to the study website. JDR is managed by the National Institute for Health Research in collaboration with the charitable organisations Alzheimer’s Society, Alzheimer’s Research UK and Alzheimer’s Scotland. It manages a list of people who have agreed to be contacted about opportunities to participate in research. The list contains over forty-five thousand people over the age of 18yrs from across the UK, and while JDR tries to actively recruit people with dementia and people who care for someone with dementia, anyone is able to sign up to the service.

The number of people on JDR eligible to take part in the current study across the UK was 47,312. The first wave of recruitment was within England from 29^th^ April and then across the rest of the UK a week later until 13^th^ May 2020 after which there was a loosening of the UK lockdown policy and therefore the decision was taken to halt data collection at that point.

The study was also advertised via Twitter and Facebook, through press releases by the University of Bristol public relations office, and through personal social media accounts. Ethical approval for the study was through the Faculty of Health Sciences Research Ethics Committee, University of Bristol (FREC ref: 103244).

### Measures

#### Sociodemographic Information

General sociodemographic information was asked of participants based on categories developed by the UK government. It included 1) general demographic details: gender, ethnicity, education, employment (fulfilling any of the UK’s key worker roles), 2) changes to employment due to COVID-19, 3) number of dependents, caring responsibilities (in particular for older adults/people with dementia), 4) smoking and alcohol consumption, 5) diagnosed sleep disorders, 6) dementia diagnosis, 7) diagnosed health conditions (heart disease, lung conditions etc.), 8) whether they are or have been infected with COVID-19.

Further items that assessed knowledge, infection, or concerns about COVID-19 were taken following World Health Organization (WHO) guidance on survey tool development for the current outbreak ^21^.

#### The Patient Health Questionnaire (PHQ8)

The Patient Health Questionnaire (PHQ8) ^22^ is an eight item self-report measure assessing symptoms of depression. It removes the ninth item from the more widely used PHQ9, “thoughts that you would be better off dead or of hurting yourself in some way” as recommended in self-administered internet surveys where further probing about positive responses to item nine are not possible ^23^. Scores range from zero to 24, with scores equal or greater than 10 indicating some level of depression being present. It has been shown to shown to be a valid diagnostic and severity measure in large clinical studies ^23^.

#### The Generalized Anxiety Disorder Questionnaire (GAD-7)

The GAD-7 is a validated self-report questionnaire that assesses generalized anxiety using 7 items with 4-point Likert scales. Total scores range from 0 to 21, with a clinical cut-offs of 5, 10 and 15 for mild, moderate and severe anxiety respectively ^24^.

#### Disordered Attitudes and Beliefs about Sleep (DBAS-16)

The DBAS-16 is a sixteen item self-report measure assessing beliefs and attitudes towards sleep using a Likert types scale ranging from 0 to 10 for items such as “I am worried that I may lose control over my abilities to sleep”. Higher scores represent more disordered attitudes around sleep. It has been shown to be a reliable and valid measure of sleep related cognitions in clinical samples^7^.

#### Assessment of sleep

The Pittsburgh Sleep Quality Index (PSQI) asks participants to report on their sleep quality and disturbances over the previous month. Nineteen individual items generate seven component scores, the sum of which gives a global score for sleep quality and disturbance. The component scores cover subjective sleep quality, sleep latency, sleep duration, habitual sleep efficiency, sleep disturbances, use of sleep medication, and daytime dysfunction. Scoring is based on a 4-point Likert-type scale ranging from 0 to 3. The scale also includes items that ask about the respondents’ bed/room partner. A global score of 5 or greater indicates a “poor” sleeper. The PSQI has good internal consistency and reliability (Cronbach’s α = .83) for its seven components^25^.

To assess the impact of the lockdown on sleep we asked participants separately whether they thought that their sleep quality had improved, stayed the same or worsened during the COVID crisis. We also presented participants with a list of 13 activities which have been reported to help people sleep, and if they had tried each activity we asked how effective they had found them at improving sleep.

#### Exercise and routine

We developed items to capture data on the number of days participants went outside during the preceding week and for how long in terms of hours and minutes. We also asked them the number of days over the previous week they had engaged in moderate to vigorous exercise (defined as activity that raises heart rate, increases breathing rate and makes them feel warmer) for 15 minutes or more. They were also asked about what time of day they went outside and engaged in exercise.

### Statistical Analysis

Descriptive statistics were used to assess participants demographic characteristics, health status, as well as mental health, attitudes towards sleep, and days outside and exercising. A One-Sample Wilcoxon test was used to assess what impact COVID-19 had on self-reported change in sleep after lockdown. We then used multinomial logistic regression to assess what factors were associated with change in self-reported sleep. To overcome differences in units of measurement across variables, we first computed z-scores for all continuous variables prior to entry into the logistic regression.

Multiple linear regression was used to assess factors associated with poor sleep in the current sample as well impact of chronotype on sleep. To assess whether sleep was worse in people with or at risk of dementia, we restricted our sample to the over 50yrs to control for age, before conducting Mood’s median test on self-reported change in sleep during lockdown, and t-test on PSQI scores. Kruskal-Wallis tests were used to assess effectiveness of sleep strategies used by participants.

An independent samples t-test was used to assess the impact of cognitive decline on sleep between those who reported they had some form of dementia or mild cognitive impairment with people from the sample over the age of 50yrs (in order to control for age).

To identify ‘larks’ and ‘owls’, for each individual we calculated the midpoint between declared wake-up and got-to-bed times. Then using a median split of this “daytime midpoint” measure, we defined individuals as ‘larks (midpoint before or at 3pm) or ‘owls’ (midpoint after 3pm). Excluding people with sleep disorders and those with no preference for time of day, we ran a fixed effects model assessing relationship between sleep quality (PSQI) and time of day outside and how that differs between ‘larks’ vs ‘owls’ controlling for age. The model was run using Maximum Likelihood Estimation. We carried a similar analysis for time of exercise (regardless of whether outside or inside).

## Results

In total, 3,474 people were included in the analysis (76% female, mean age 59.2 years (Sd. 13.3) – Table 1). Forty-six people with any form of dementia and 33 with MCI provided data.

**Table 1:**
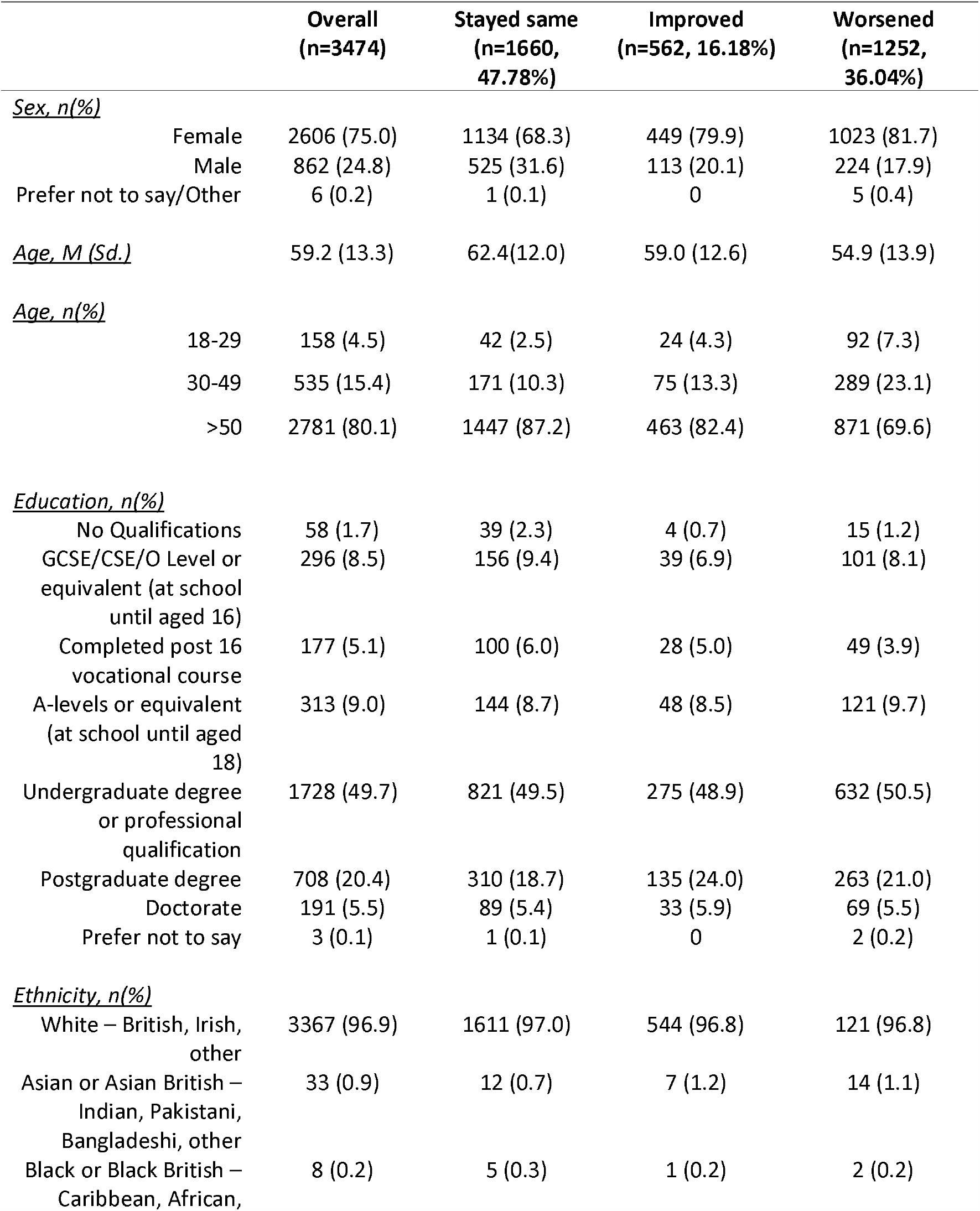
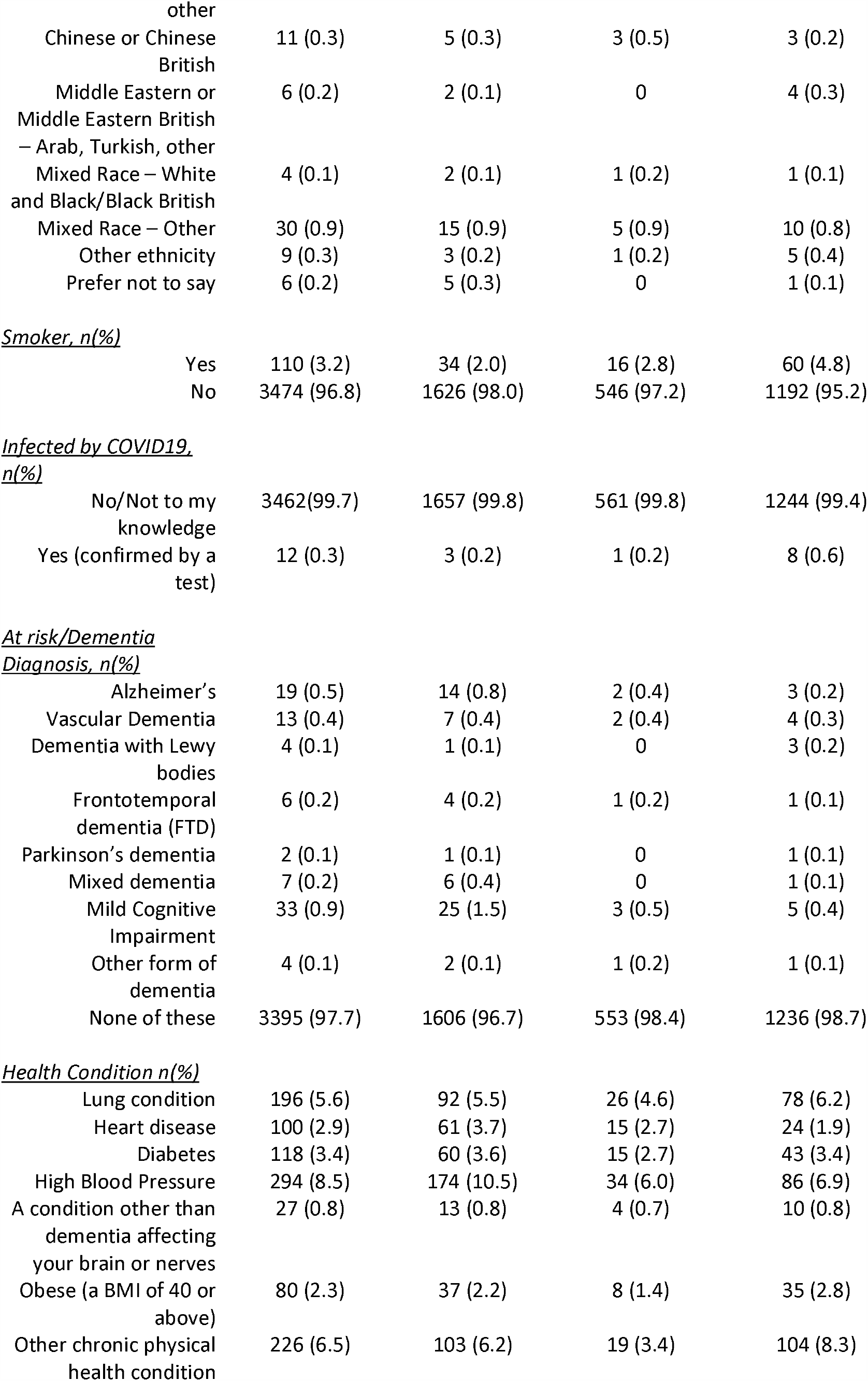
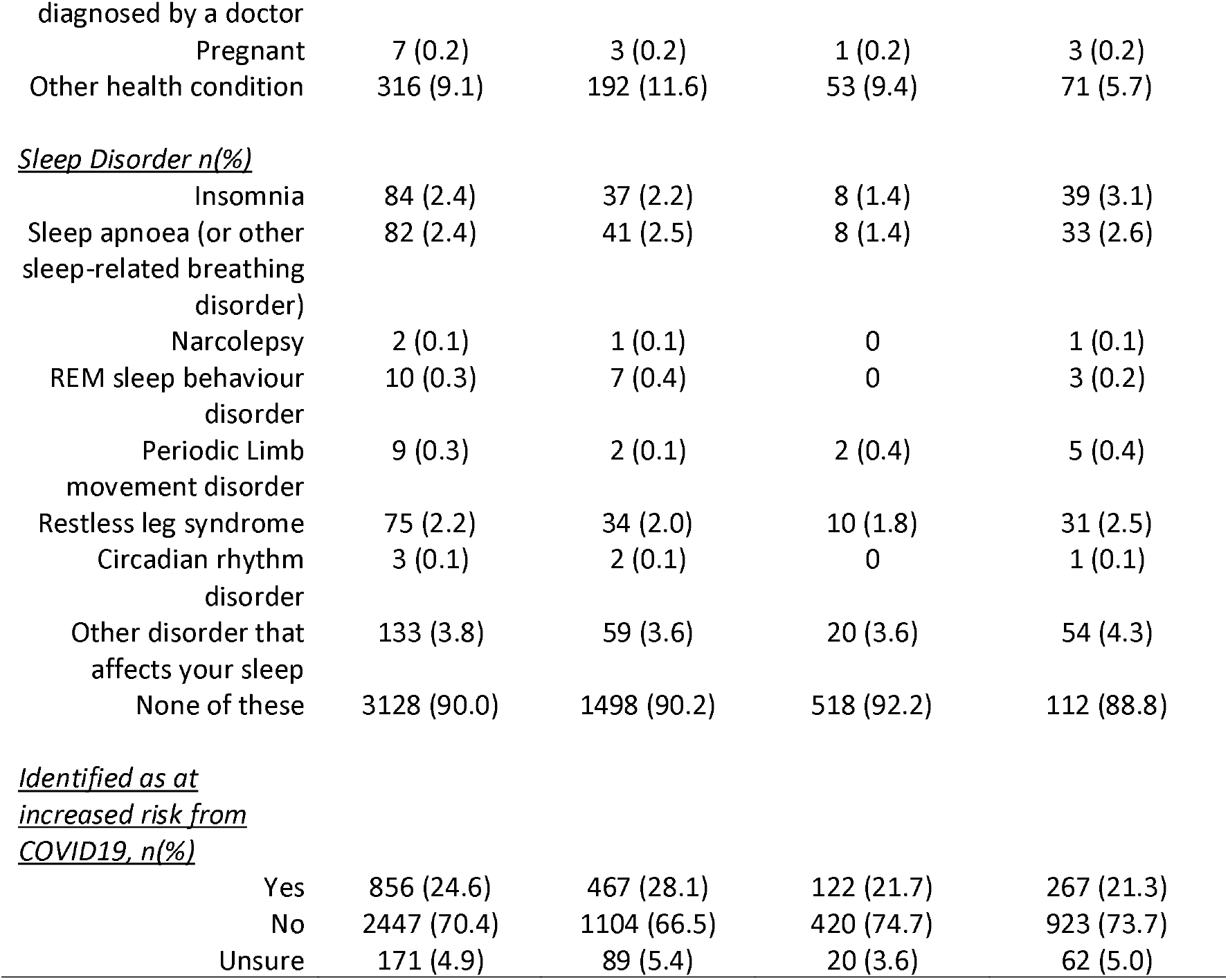
Demographic details for the complete sample and split between those who reported their sleep staying the same, improving or worsening.

|  |  | **Overall**<br>**(n=3474)** | **Stayed same**<br>**(n=1660,**<br>**47.78%)** | **Improved**<br>**(n=562, 16.18%)** | **Worsened**<br>**(n=1252,**<br>**36.04%)** |
| --- | --- | --- | --- | --- | --- |
| <u>*Sex, n(%)*</u> |  |  |  |  |  |
|  | Female | 2606 (75.0) | 1134 (68.3) | 449 (79.9) | 1023 (81.7) |
|  | Male | 862 (24.8) | 525 (31.6) | 113 (20.1) | 224 (17.9) |
|  | Prefer not to say/Other | 6 (0.2) | 1 (0.1) | 0 | 5 (0.4) |
| <u>*Age, M (Sd.)*</u> |  | 59.2 (13.3) | 62.4 (12.0) | 59.0 (12.6) | 54.9 (13.9) |
| <u>*Age, n(%)*</u> |  |  |  |  |  |
|  | 18-29 | 158 (4.5) | 42 (2.5) | 24 (4.3) | 92 (7.3) |
|  | 30-49 | 535 (15.4) | 171 (10.3) | 75 (13.3) | 289 (23.1) |
|  | >50 | 2781 (80.1) | 1447 (87.2) | 463 (82.4) | 871 (69.6) |
| <u>*Education, n(%)*</u> |  |  |  |  |  |
|  | No Qualifications | 58 (1.7) | 39 (2.3) | 4 (0.7) | 15 (1.2) |
|  | GCSE/CSE/O Level or equivalent (at school until aged 16) | 296 (8.5) | 156 (9.4) | 39 (6.9) | 101 (8.1) |
|  | Completed post 16 vocational course | 177 (5.1) | 100 (6.0) | 28 (5.0) | 49 (3.9) |
|  | A-levels or equivalent (at school until aged 18) | 313 (9.0) | 144 (8.7) | 48 (8.5) | 121 (9.7) |
|  | Undergraduate degree or professional qualification | 1728 (49.7) | 821 (49.5) | 275 (48.9) | 632 (50.5) |
|  | Postgraduate degree | 708 (20.4) | 310 (18.7) | 135 (24.0) | 263 (21.0) |
|  | Doctorate | 191 (5.5) | 89 (5.4) | 33 (5.9) | 69 (5.5) |
|  | Prefer not to say | 3 (0.1) | 1 (0.1) | 0 | 2 (0.2) |
| <u>*Ethnicity, n(%)*</u> |  |  |  |  |  |
|  | White – British, Irish, other | 3367 (96.9) | 1611 (97.0) | 544 (96.8) | 121 (96.8) |
|  | Asian or Asian British – Indian, Pakistani, Bangladeshi, other | 33 (0.9) | 12 (0.7) | 7 (1.2) | 14 (1.1) |
|  | Black or Black British – Caribbean, African, | 8 (0.2) | 5 (0.3) | 1 (0.2) | 2 (0.2) |

|  |  |  |  |  |
| --- | --- | --- | --- | --- |
| other |  |  |  |  |
| Chinese or Chinese British | 11 (0.3) | 5 (0.3) | 3 (0.5) | 3 (0.2) |
| Middle Eastern or Middle Eastern British – Arab, Turkish, other | 6 (0.2) | 2 (0.1) | 0 | 4 (0.3) |
| Mixed Race – White and Black/Black British | 4 (0.1) | 2 (0.1) | 1 (0.2) | 1 (0.1) |
| Mixed Race – Other | 30 (0.9) | 15 (0.9) | 5 (0.9) | 10 (0.8) |
| Other ethnicity | 9 (0.3) | 3 (0.2) | 1 (0.2) | 5 (0.4) |
| Prefer not to say | 6 (0.2) | 5 (0.3) | 0 | 1 (0.1) |
| <u>Smoker, n(%)</u> |  |  |  |  |
| Yes | 110 (3.2) | 34 (2.0) | 16 (2.8) | 60 (4.8) |
| No | 3474 (96.8) | 1626 (98.0) | 546 (97.2) | 1192 (95.2) |
| <u>Infected by COVID19, n(%)</u> |  |  |  |  |
| No/Not to my knowledge | 3462(99.7) | 1657 (99.8) | 561 (99.8) | 1244 (99.4) |
| Yes (confirmed by a test) | 12 (0.3) | 3 (0.2) | 1 (0.2) | 8 (0.6) |
| <u>At risk/Dementia Diagnosis, n(%)</u> |  |  |  |  |
| Alzheimer's | 19 (0.5) | 14 (0.8) | 2 (0.4) | 3 (0.2) |
| Vascular Dementia | 13 (0.4) | 7 (0.4) | 2 (0.4) | 4 (0.3) |
| Dementia with Lewy bodies | 4 (0.1) | 1 (0.1) | 0 | 3 (0.2) |
| Frontotemporal dementia (FTD) | 6 (0.2) | 4 (0.2) | 1 (0.2) | 1 (0.1) |
| Parkinson's dementia | 2 (0.1) | 1 (0.1) | 0 | 1 (0.1) |
| Mixed dementia | 7 (0.2) | 6 (0.4) | 0 | 1 (0.1) |
| Mild Cognitive Impairment | 33 (0.9) | 25 (1.5) | 3 (0.5) | 5 (0.4) |
| Other form of dementia | 4 (0.1) | 2 (0.1) | 1 (0.2) | 1 (0.1) |
| None of these | 3395 (97.7) | 1606 (96.7) | 553 (98.4) | 1236 (98.7) |
| <u>Health Condition n(%)</u> |  |  |  |  |
| Lung condition | 196 (5.6) | 92 (5.5) | 26 (4.6) | 78 (6.2) |
| Heart disease | 100 (2.9) | 61 (3.7) | 15 (2.7) | 24 (1.9) |
| Diabetes | 118 (3.4) | 60 (3.6) | 15 (2.7) | 43 (3.4) |
| High Blood Pressure | 294 (8.5) | 174 (10.5) | 34 (6.0) | 86 (6.9) |
| A condition other than dementia affecting your brain or nerves | 27 (0.8) | 13 (0.8) | 4 (0.7) | 10 (0.8) |
| Obese (a BMI of 40 or above) | 80 (2.3) | 37 (2.2) | 8 (1.4) | 35 (2.8) |
| Other chronic physical health condition | 226 (6.5) | 103 (6.2) | 19 (3.4) | 104 (8.3) |
| diagnosed by a doctor |  |  |  |  |
| Pregnant | 7 (0.2) | 3 (0.2) | 1 (0.2) | 3 (0.2) |
| Other health condition | 316 (9.1) | 192 (11.6) | 53 (9.4) | 71 (5.7) |
| *<u>Sleep Disorder n(%)</u>* |  |  |  |  |
| Insomnia | 84 (2.4) | 37 (2.2) | 8 (1.4) | 39 (3.1) |
| Sleep apnoea (or other sleep-related breathing disorder) | 82 (2.4) | 41 (2.5) | 8 (1.4) | 33 (2.6) |
| Narcolepsy | 2 (0.1) | 1 (0.1) | 0 | 1 (0.1) |
| REM sleep behaviour disorder | 10 (0.3) | 7 (0.4) | 0 | 3 (0.2) |
| Periodic Limb movement disorder | 9 (0.3) | 2 (0.1) | 2 (0.4) | 5 (0.4) |
| Restless leg syndrome | 75 (2.2) | 34 (2.0) | 10 (1.8) | 31 (2.5) |
| Circadian rhythm disorder | 3 (0.1) | 2 (0.1) | 0 | 1 (0.1) |
| Other disorder that affects your sleep | 133 (3.8) | 59 (3.6) | 20 (3.6) | 54 (4.3) |
| None of these | 3128 (90.0) | 1498 (90.2) | 518 (92.2) | 112 (88.8) |
| *<u>Identified as at increased risk from COVID19, n(%)</u>* |  |  |  |  |
| Yes | 856 (24.6) | 467 (28.1) | 122 (21.7) | 267 (21.3) |
| No | 2447 (70.4) | 1104 (66.5) | 420 (74.7) | 923 (73.7) |
| Unsure | 171 (4.9) | 89 (5.4) | 20 (3.6) | 62 (5.0) |

### Lockdown has impaired sleep quality overall

More participants stated that their sleep had deteriorated (1,252, 47.78%) as opposed to improved (562, 16.18%) since the start of the UK lockdown, with 1,660 (47.78%) reporting that their sleep had stayed the same (Figure 1A). One-Sample Wilcoxon Test provides evidence that the sample median differed from the expected value (“Stayed the same”) (p< 0.001) suggesting that, overall, lockdown has negatively affected sleep quality. However, it is also relevant that lockdown has improved sleep for some. In the next sections we explore markers of perceived change in sleep and overall sleep quality (PSQI total score) during lockdown with a focus on identifying potentially modifiable risk and protective factors.

**Figure 1.**
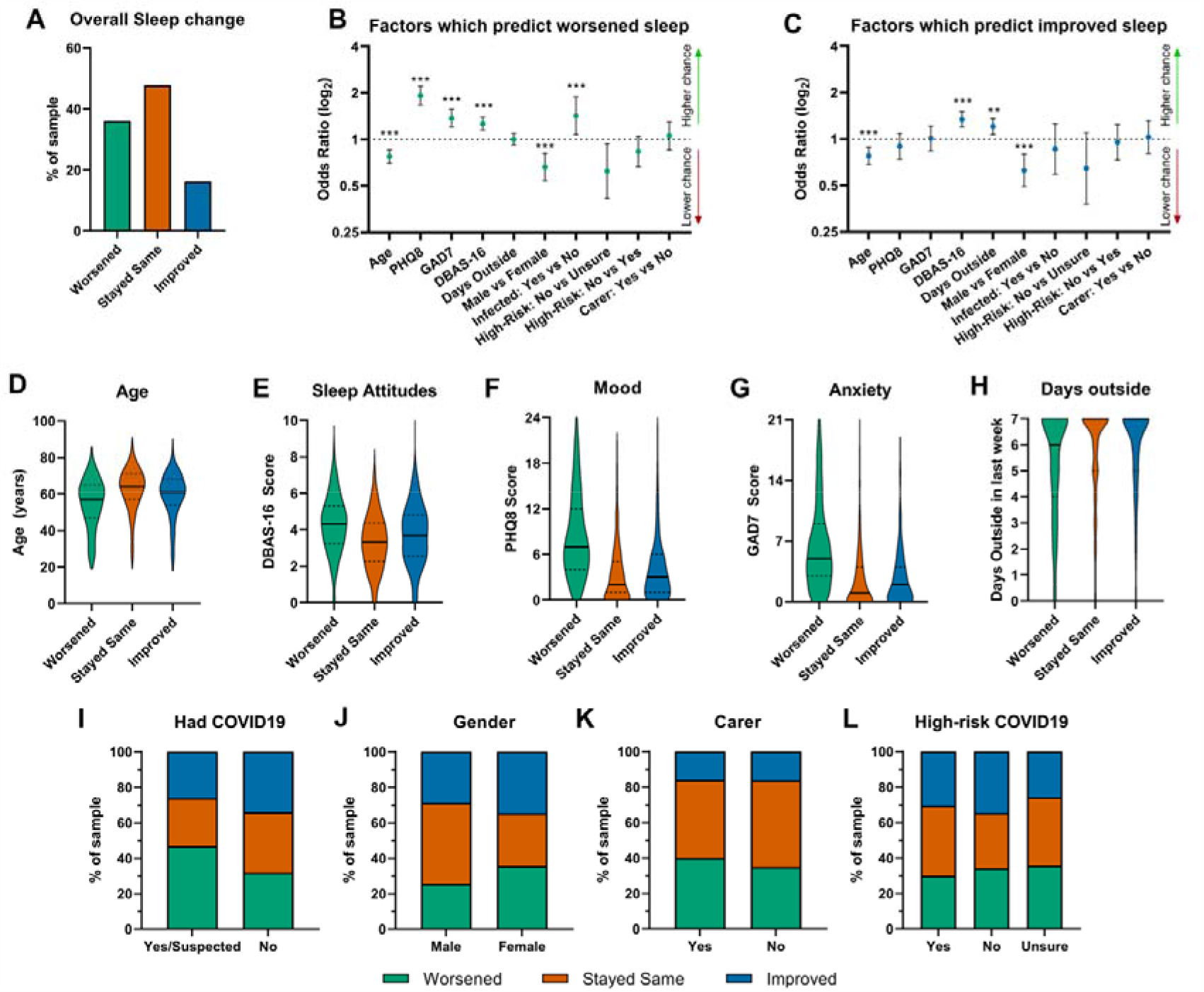
Factors affecting sleep change during lockdown. (A) Sleep change across the entire cohort. (B-C) Summary results (odds ratios ± 95% CIs) from multinomial logistic regression predicting sleep change. Significant predictors are marked with asterisks (**p< 0.01 ***p< 0.001). Predictors are shown separately as to whether they predict improved (B) or worsened sleep (C). Y axes are scaled by log2. Each predictor is individually shown in panels D-L. Continuous variable predictors are shown as violin plots (D-H). Solid black lines represent the median. Categorical variable predictors are shown in panels I-L, in which each bar represents all individuals within a given category, split by proportions of sleep change.

### Anxiety, depression, and disordered attitudes were associated with sleep worsening, whereas time outside was associated with improvement of sleep during lockdown (Figure 1; Table 2)

First, we investigated which factors were associated with self-report of sleep improving or deteriorating (compared to staying the same). There was evidence of a positive association between level of anxiety and lockdown-induced worsening of sleep (one standard deviation change in GAD7 – OR=1.37; 95% CI=[1.20, 1.57]). Similarly, for mood, there was evidence that increase in mood score (standard deviation change in PHQ8) was associated with worsening of sleep during lockdown (OR=1.92; 95% CI=[1.67, 2.21]). There was evidence that COVID infection (OR=1.42; 95% CI=[1.07, 1.88]) and perceived risk of COVID (OR=0.62; 95% CI=[0.41, 0.94]) were both associated with reported worsened sleep.

**Table 2:** Multinomial Logistic Regression of standardised variables with sleep “Stayed the Same” as the outcome reference category (overall total n=3,362). Data shown graphically in Figure 1 B & C

|  | <u>Worsened</u> |  | <u>Improved</u> |  |
| --- | --- | --- | --- | --- |
|  | <u>OR</u> | <u>(95% CI)</u> | <u>OR</u> | <u>(95% CI)</u> |
| Age | 0.78 | (0.70, 0.86) *** | 0.78 | (0.69, 0.88) *** |
| PHQ8 | 1.92 | (1.67, 2.21) *** | 0.90 | (0.74, 1.09) |
| GAD7 | 1.37 | (1.20, 1.57) *** | 1.01 | (0.84, 1.22) |
| DBAS-16 | 1.26 | (1.14, 1.39) *** | 1.34 | (1.19, 1.51) *** |
| Days outside | 1.00 | (0.92, 1.10) | 1.20 | (1.07, 1.36) ** |
| <u>Gender</u> |  |  |  |  |
| Male | 0.66 | (0.54, 0.81) *** | 0.63 | (0.49, 0.80) *** |
| Female | ref |  | ref |  |
| <u>Infected with COVID19</u> |  |  |  |  |
| Yes | 1.42 | (1.07, 1.88) * | 0.86 | (0.59, 1.26) |
| No | ref |  | ref |  |
| <u>Increased risk from COVID19</u> |  |  |  |  |
| Unsure | 0.62 | (0.41, 0.94) * | 0.65 | (0.38, 1.10) |
| Yes | 0.83 | (0.67, 1.05) | 0.95 | (0.73, 1.24) |
| No | ref |  | ref |  |
| <u>Caring responsibilities</u> |  |  |  |  |
| Yes | 1.05 | (0.86, 1.30) | 1.03 | (0.81, 1.32) |
| No | ref |  | ref |  |

Abbreviations: GAD, generalized anxiety disorder; PHQ, patient health questionnaire; DBAS, Disordered Beliefs and Attitudes to Sleep; OR, odds ratio; 95% CI, 95% confidence interval; *ref*, reference category; COVID-19, 2019 Corona Virus Disease. \* $P < 0.05$ , \*\* $P < 0.01$ , \*\*\* $P < 0.001$

In contrast, there was evidence that higher number of days outside in the preceding week was associated with an increase in the odds that that sleep would be reported to have improved vs stayed the same (OR=1.20; 95% CI=[1.07, 1.36]).

Two factors appeared to make sleep more resistant to lock down-induced change: increasing age (standard deviation change in age) was associated with reduced odds of sleep improving (OR=0.78; 95% CI=[0.69, 0.88]) or worsening (OR=0.78; 95% CI=[0.70, 0.86]) versus stayed the same, and being male reduced odds of sleep being reported as having improved versus stayed the same (OR=0.63; 95% CI=[0.49, 0.80]) as well as deteriorating versus staying the same (OR=0.66; 95% CI=[0.54, 0.81]). In contrast, more disordered beliefs and attitudes towards sleep (DBAS-16), was associated with greater likelihood of reported sleep improving compared to staying the same (OR=1.34; 95% CI=[1.19, 1.51]) as well as sleep worsening compared to staying the same (OR=1.26; 95% CI=[1.14, 1.39]).

We also used PSQI as a more objective measure of sleep quality over the preceding month (all within lockdown). There was evidence that age, whether they were female, dysfunctional attitudes to sleep, depression and anxiety, were associated with poorer sleep quality, while not being infected by COVID-19 and not having caring responsibilities were associated with improved sleep (F_9,3349_ = 259.45, p < 0.001; adjusted R^2^ = 0.41) (see Table 3).

**Table 3:** Multiple linear regression for variables associated with PSQI during COVID19 lockdown.

|  | <u>B</u> | <u>(95% CI)</u> | <u>Beta</u> | <u>t</u> |  |
| --- | --- | --- | --- | --- | --- |
| Age | 0.03 | (0.02, 0.03) | 0.12 | 7.87 | *** |
| Female | 0.45 | (0.26, 0.63) | 0.07 | 4.78 | *** |
| DBAS-16 | 0.35 | (0.29, 0.41) | 0.18 | 12.09 | *** |
| GAD7 | 0.06 | (0.03, 0.08) | 0.08 | 3.96 | *** |
| PHQ8 | 0.28 | (0.25, 0.30) | 0.48 | 21.98 | *** |
| Days outdoors | -0.01 | (-0.06, 0.04) | -0.01 | -0.38 |  |
| Increased risk from COVID19 | -0.05 | (-0.20, 0.10) | -0.01 | -0.66 |  |
| Infected by COVID19 | -0.31 | (-0.57, -0.04) | -0.03 | -2.29 | * |
| No caring responsibilities | -0.29 | (-0.48, -0.10) | -0.04 | -2.94 | ** |

Abbreviations: PSQI, Pittsburgh Sleep Quality Index; GAD, generalized anxiety disorder; PHQ, patient health questionnaire; DBAS, Disordered Beliefs and Attitudes to Sleep; CI, confidence interval for B; COVID-19, 2019 Corona Virus Disease. \* $P < 0.05$ , \*\* $P < 0.01$ , \*\*\* $P < 0.001$ Adjusted $R^2 = 0.41$ , ( $df=9$ , 3349, $p < 0.001$ )

### Poorer sleep in people with dementia and their carers (Figure 2)

Of the 76 people who responded with a self-reported diagnosis of dementia or MCI, 54 (71.05%) stated that their sleep stayed the same, 9 (11.84%) improved and for 13 (17.11%) their sleep got worse. To more closely age-match groups, only people over 50 were included in the following analysis. An independent samples t-test in over 50s comparing people with dementia/MCI with those without suggested higher PSQI score (worse sleep) in the dementia/MCI group (Mean Diff = 0.74; 95% CI [0.06, 1.44]). Interestingly, 31.7% of people without dementia more often reported sleep worsening during lockdown compared to 17.1% of those with dementia/MCI (p < 0.001).

**Figure 2.**
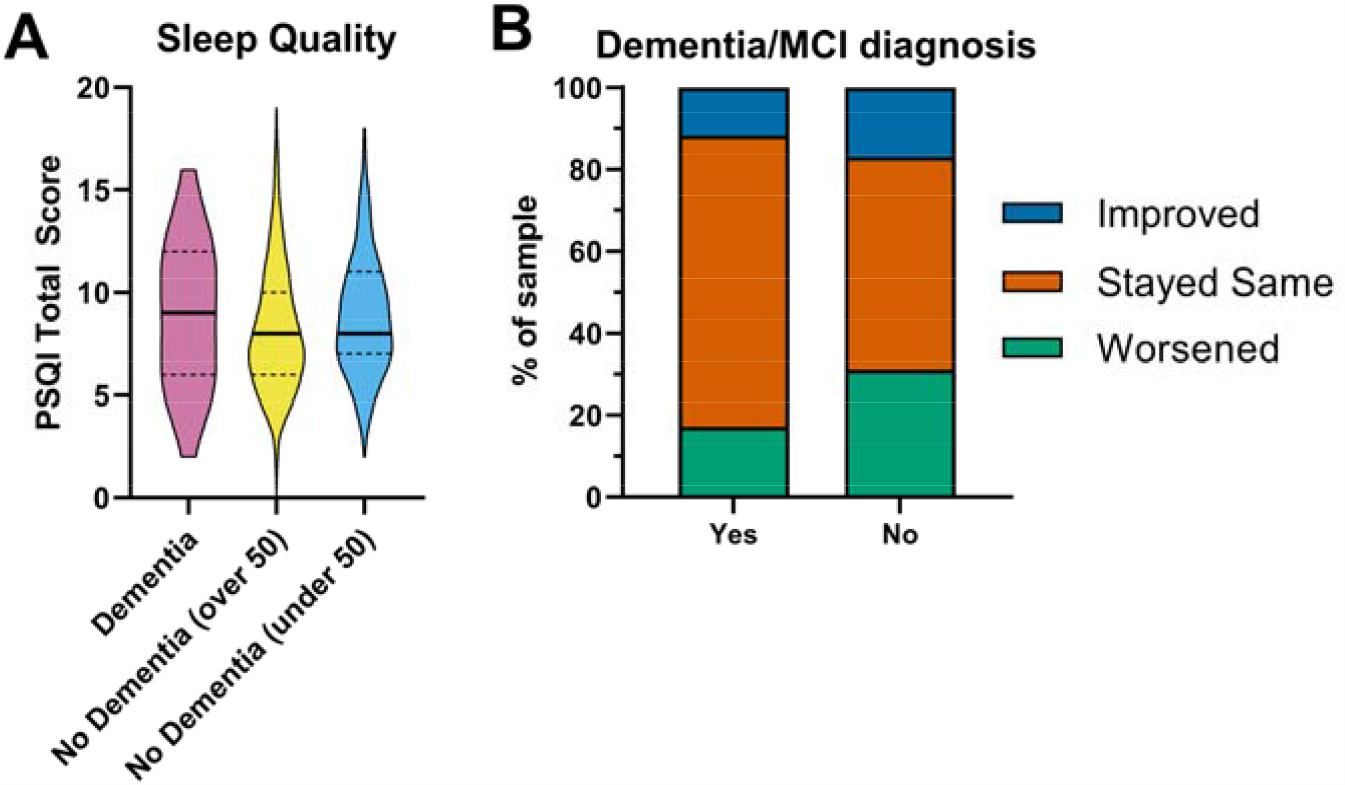
How sleep affects people with Dementia and Mild Cognitive Impairment. Absolute Sleep quality as measured by PSQI is shown in panel A. Panel B shows which proportions of each group that have reported sleep change, or lack thereof.

For people with caring responsibilities, their sleep was generally worse compared to those who did not care for someone (Mean PSQI Diff = 0.70; 95% CI [0.46, 0.94]), and they reported that their sleep had also worsened to a greater extent that those without caring responsibilities (40.2% vs. 34.8%; p < 0.01).

### Self-help with sleep – what works?

All strategies were found to be at least somewhat useful, with positive effectiveness scores (ES), however medians of each activity-type did differ significantly (K-W statistic = 2591, p<0.001; Figure 3). “Doing exercise” during the day was reported to aid sleep the most (ES = 1.21, 95% CI=[1.18, 1.24], n=3476); all other strategies were therefore statistically compared to this using Dunn’s test for pairwise comparisons. Reducing caffeine before bed was the only strategy that did not appear to be less effective than daytime exercise (ES = 1.15, 95% CI=[1.11,1.18], n=2906, p_Dunn_=0.334). Keeping a routine (which may have included exercise) was the next most effective strategy (ES = 1.14, 95% CI=[1.11, 1.18], n=3339, p_Dunn_=0.048) followed by reading before bed (ES = 1.11, 95% CI=[1.08, 1.15], n=3179, p=0.0005). All other strategies were significantly less effective than daytime exercise at the p<.0001 level. The least effective reported strategy was having a hot drink before bed (ES = 0.25, 95% CI=[0.21, 0.29], n=2384, p<.0001).

**Figure 3.**
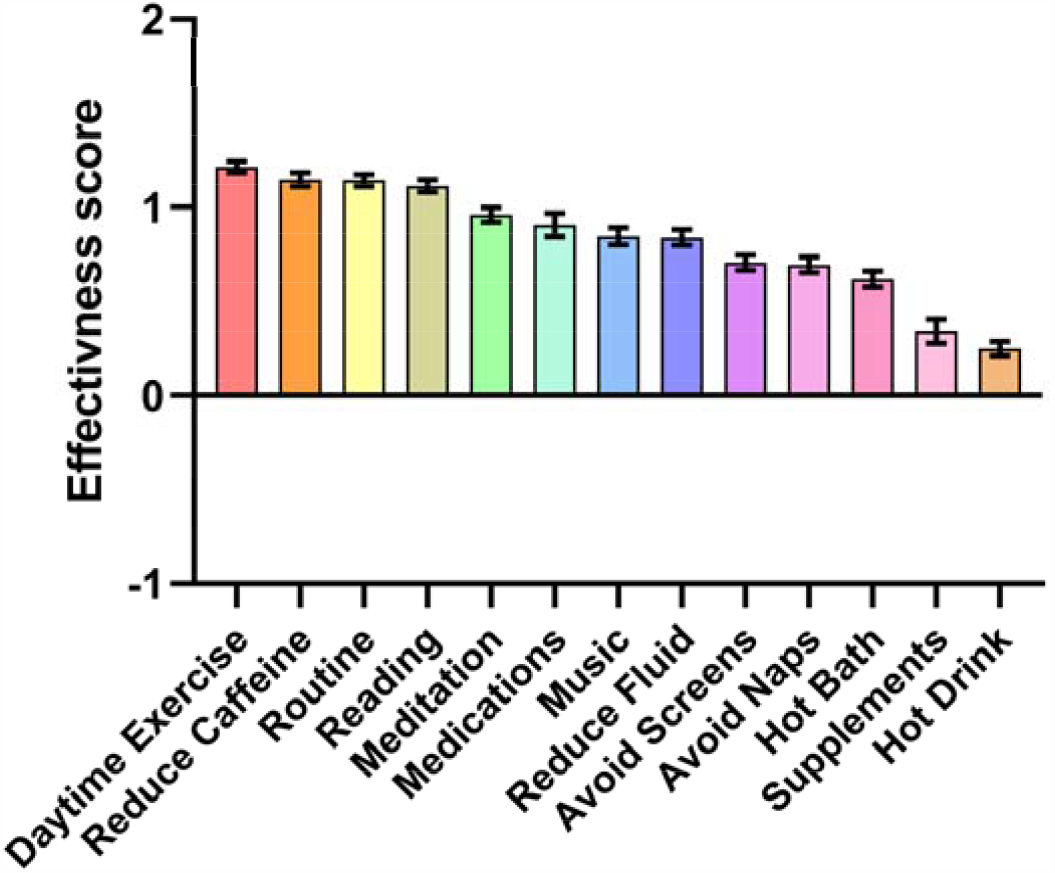
Perceived effectiveness of different sleep strategies. Strategies are ranked in order of reported effectiveness (left-right). Y axis shows total possible score range (−1—+2).

In summary, the most effective self-help strategies to enhance sleep are daytime exercise, sticking to a daily routine, avoiding caffeine before bed, and reading at bedtime.

### Time of day being outside, but not time of day of exercise, predicts sleep quality differently for ‘larks’ and ‘owls’

Given amount of time spent outside was a factor that appeared to improve sleep, we explored further whether time of day of outside impacted sleep quality. Of 2065 people who declared a typical time of day for being outside over the preceding week, time of day of being outside predicts change in sleep with being outside before midday protecting against worsening of sleep (Before 9am: OR = 1.99 95% CI =[1.21, 3.26], 9am-midday: OR = 1.78 95% CI=[1.15, 2.75]; both with ‘After 6pm’ as reference category) whereas being outside at other times of day had no effect on change in sleep after correcting for age. Similarly, PSQI was predicted by time of day outside, with being outside earlier in the day predicting better sleep (lower PSQI) – (F(2,2063)=11.77, p<.001, β= 0.057, p=.010).

We next asked whether chronotype affected the relationship between time of day of being outside and sleep. We showed that PSQI in both groups was better for those who spent more time outdoors (‘larks: F(2,1687)=13.63, p<.001, β=-0.90, p=.005, ‘owls’: F(2,1760)=32.39, p<.001, β= −0.19, p<.001). However, there was a dissociation in the effect of time of day of being outside between ‘larks and ‘owls’. In a model to predict sleep quality in ‘larks’ and ‘owls’, we showed age was a significant predictor (F(1,1874)=20.33, p<.001) of sleep quality and even after controlling for age, there was an interaction between chronotype and time of day outside (F(4,1874)=4.214, p=.040; Figure 4a). We explored this further and demonstrated a linear relationship between PSQI and time outside for ‘owls’ (Linear: F(1,1050) = 13.99, p<.001, Non-Linear F(3,1050) = 0.251, p=.861) and a non-linear relationship for ‘larks (Linear: F(1, 1008) = 1.20, p=.274), Non-Linear: F(3,1008) = 2.77, p=.041).

**Figure 4.**
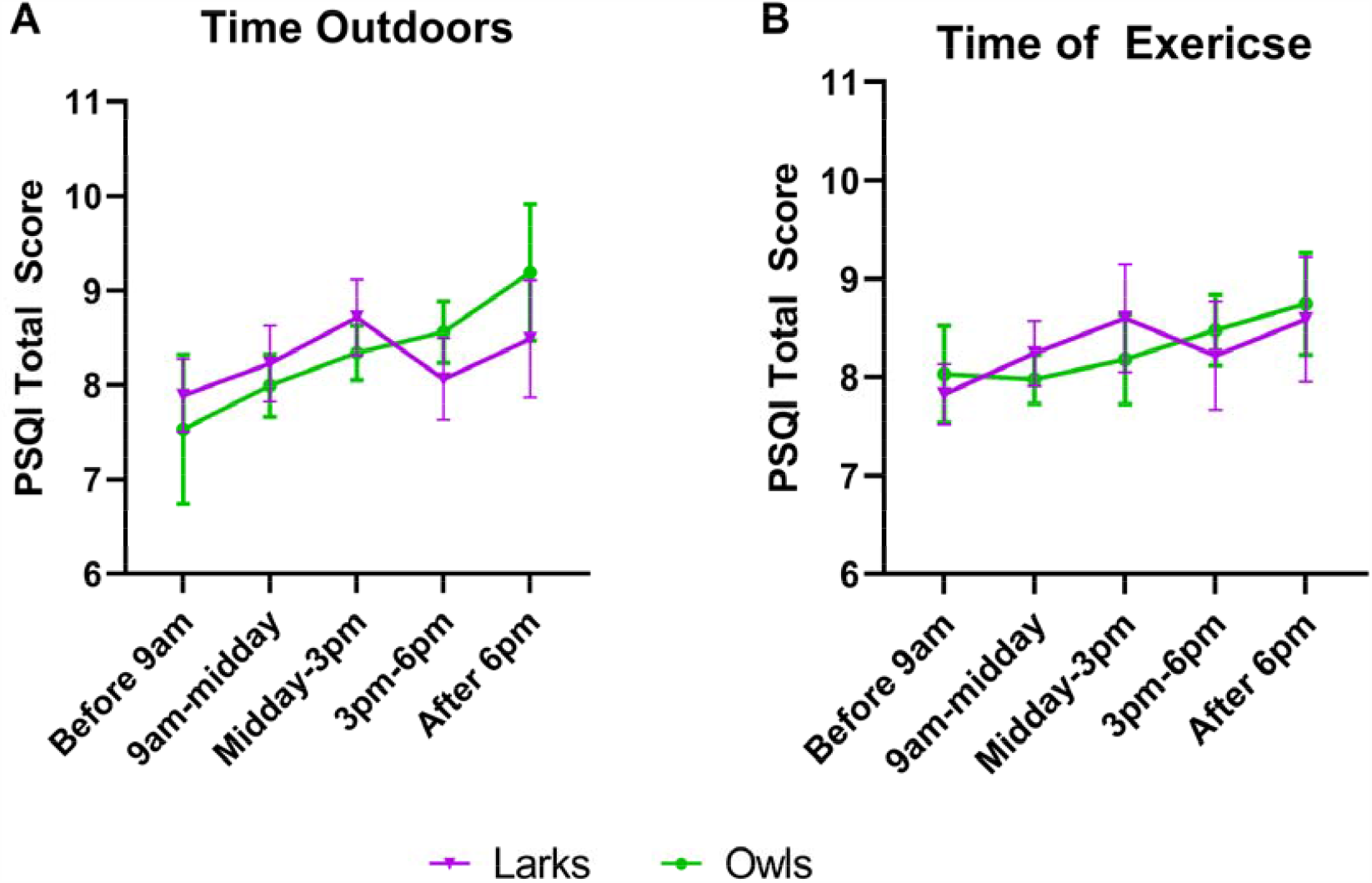
Effect of daily routine on sleep quality of people of different chronotypes (‘Lark’ and ‘Owl’). People were asked what time, if any, they typically spent outdoors (A) or doing exercise (B). Error bars show mean ± 95% CI for each time option. People with no typical time were excluded from this analysis. Higher PSQI scores represent poorer sleep quality.

Time of day of being outside during lockdown was understandably highly correlated with time of day of exercise (Pearson R =0.654, p<.001) and therefore we included only time of day of being outside in the main model described above. When we look separately at exercise a predictor of sleep, time of day of exercise also predicted sleep (after controlling for age) – F(1,1901)=8.298, p=.004. Interestingly, there was no interaction between time of day of exercise (regardless of whether outside or not) and chronotype in predicting sleep quality (F(4,1901)=0.268, p=.605; Figure 4b). This suggests, perhaps, that being outside, rather than physical activity, is the key factor that differentiates the relationship between time of day of activity and sleep quality in ‘larks’ and ‘owls’. Overall, we clearly show a benefit of early exercise and outside activity, and time of day of being outside and exercise, on sleep quality overall.

## Discussion

Here we show evidence that overall COVID-19 lockdown has impaired sleep in the UK in line with the international picture^11 10^. Factors associated with reported improved sleep during lockdown include spending more time outside. Factors that were associated with subjective worsening of sleep included higher levels of depression and anxiety and infection with COVID-19 or suspected infection. Interestingly, older age and being male was associated with a tendency for sleep to stay the same and Dysfunctional Attitudes to Sleep resulted in higher levels of both improving and deteriorating sleep, suggesting ageing makes sleep more resilient to change, whereas, attitude to sleep can make it more susceptible to change. Poor sleep during lockdown as measured by the validated PSQI measure was associated with caring responsibility, depression, anxiety, female gender, COVID infection and dysfunctional attitude to sleep.

Taking advantage of the unique opportunity to study very regulated trips outside the home for exercise (once per day stipulated by UK government), we also demonstrate that being outside and exercise earlier in the day are associated with better sleep. However, the benefit of being outside early was present only for people with later chronotypes (‘owls’) and not people with early chronotypes (‘larks’). Although, time outside and exercise were highly correlated, people did have the opportunity to exercise inside and could be outside in gardens without significant exercise. Exercise alone (regardless of whether outside or not), promoted sleep, but there was no dissociation between ‘larks’ and ‘owls’ in the effect of early exercise on sleep quality.

### Lockdown impairment of sleep is associated with depression, anxiety

In keeping with a role for depression, stress and anxiety in poor sleep ^5^, measures of anxiety and depression were significant predictors of worsening of sleep following the onset of lockdown conditions. It is unclear from our data whether COVID-19 lockdown caused the higher rates of sleep difficulties in our sample, or whether pre-existing anxiety and depression led to an increased susceptibility to worsening of sleep. However, our findings are in line with other research showing links between the COVID-19 outbreak and higher levels of anxiety, low mood and sleeplessness in other countries ^10,11^.

A previous study during the 2007 equine influenza outbreak in Australia, where movement was restricted and households were quarantined, found that younger people experienced the most psychological distress ^26^ which might explain why sleep is more affected in young than older people in our study.

### Attitude to sleep

As expected, we found that dysfunctional beliefs and attitudes towards sleep predicted higher levels of disordered sleep. Previous research has highlighted the role that unhelpful beliefs and attitudes towards sleep play in maintaining insomnia ^27^. People who experience difficulties sleeping may hold unrealistic expectations about how much sleep they need and then worry if they are not achieving this. This may be because, for example, their perceived impact on future daytime functioning, or concerns about the possible health impact of poor sleep. These unhelpful beliefs and cognitive arousal lead to physiological arousal, which in turn further hampers achieving good quality sleep and maintaining a vicious cycle of insomnia ^7^. Much has been made in the media recently about the negative impact of COVID-19 on sleep ^9^ which is likely to further concerns about sleep quality in the general population. Our findings confirm that people who said their sleep had worsened since the start of lockdown, had higher rates of disordered beliefs and attitudes towards sleep which may have contributed to the worsening of sleep due to lockdown.

### Timing of physical activity or daylight exposure to improve sleep

COVID lockdown restrictions in the UK at the time of the questionnaire dictated that people could only leave their house once per day for exercise. In line with our expectations, higher number of days outside in the preceding week predicted better sleep during lockdown ^28^. Being outside and exercising early in the day were associated with better sleep ^29^. However, when we explored this further, we found that chronotype moderated this association for being outside only – being outside early improved sleep quality only in people with a late or ‘owl’ chronotypes. There was no such interaction for time of day of exercise (regardless of whether inside or outside). Therefore, it is likely that daylight early in the day is linked to better sleep for people who tend towards a later chronotype – plausibly through advancing circadian phase more for ‘owls’ than ‘larks’ or through a direct effect on sleep. Advancing the circadian clock through exercise and light exposure could better align sleep time with endogenous hormonal rhythmicity ^17,30^ and/or environmental factors conducive to sleep, for example, darkness and quiet. Furthermore, exercise and daylight exposure can have a direct effect on mood which in turn can improve sleep quality.

Early exercise and light exposure as part of an intervention have previously resulted in circadian advance and improved wellbeing in a small group of young people^31^. Here we show potentially for such intervention in older people, where sleep is generally poorer, as our finding was true for the whole group after taking account of age.

### Limitations

Our main limitation is its cross-sectional design that prohibits causal inference about why sleep has deteriorated other than participants’ self-report. This is inevitable as data were collected after the outbreak of COVID-19 and we were unable to assess participants’ level of psychological distress or sleep prior to the lockdown being imposed. The sample was drawn overwhelmingly from people who has signed-up to JDR and were happy to volunteer for an online questionnaire. Hence the findings should be considered in the light of possible selection bias. Assessment of chronotype was very simply done – midpoint of wake and sleep time – and our measures of exercise will include unmeasured confounds such as different types of exercise in ‘owls’ and ‘larks’. However, we expect that these issues have introduced noise (which is to some extent overcome by large sample size) rather than a systematic bias.

## Conclusion

Many of the factors associated with lockdown-related sleep change are modifiable, for example, anxiety, depression and time of day of being outside and exercise. Effective interventions should be tailored to chronotype. While older people have poorer sleep than younger people, the impact of COVID-19 lockdown on the sleep was greater in younger people. Sleep problems entrenched during youth can persist, even after the initial stressor had resolved, due to a number of behavioural and psychological factors. Therefore, managing sleep disorders across the age range will help to protect population brain health in people who have lived through the pandemic. Here, we share strategies employed by our participants that are thought to have helped sleep and provide the foundation for development of future personalised interventions.

## Data Availability

Fully anonymised data will be available to bona fide researchers on request as guided by university of Bristol data access policy?

## Acknowledgments

This work was funded by a Above & Beyond grant (ABL-2019-20-01) and by Alzheimer’s Research UK (ARUK-NC2019-BB). Special thanks go to James Grassom and Adam Smith at Join Dementia Research for their help in placing the study on the JDR platform and supporting the recruitment and advertising for this study. Also, thanks go to Joanne Fryer in the Media Team at the University of Bristol Press Office for issuing a press release to advertise the study. Finally, we would also like to thank Marta Swirski as well as input from the Researching Sleep to Treat Dementia (RESTED) stakeholder group and the Patient and Public Involvement group in helping us to develop the research questions and methods reported here.

## Author contributions

Conceptualisation: N.C, A.W, E.C.; Formal Analysis: N.C, A.W, E.C., N.T., with help from J.S.; Writing – Original Draft: N.C, A.W, E.C with help from S.M; Writing – Reviewing & Editing: N.C, A.W, E.C., S.M, H.P., N.T.

## References

1. Besedovsky L, Lange T, Haack M. The Sleep-Immune Crosstalk in Health and Disease. Physiol Rev. 2019;99(3):1325–1380.

2. St-Onge M-P, Zuraikat FM. Reciprocal roles of sleep and diet in cardiovascular health: a review of recent evidence and a potential mechanism. Current atherosclerosis reports. 2019;21(3):11.

3. Reid KJ, Matinovich Z, Finkel S, et al. Sleep: A Marker of Physical and Mental Health in the Elderly. American Journal of Geriatric Psychiatry. 2006;14:860–866.

4. Tranah GJ, Blackwell T, Stone KL, et al. Circadian activity rhythms and risk of incident dementia and mild cognitive impairment in older women. Annals of neurology. 2011;70(5):722–732.

5. Gross RT, Borkovec T. Effects of a cognitive intrusion manipulation on the sleep-onset latency of good sleepers. Behavior Therapy. 1982;13(1):112–116.

6. Nau SD, McCrae CS, Cook KG, Lichstein KL. Treatment of insomnia in older adults. Clinical psychology review. 2005;25(5):645–672.

7. Morin CM, Vallieres A, Ivers H. Dysfunctional Beliefs and Attitudes about Sleep (DBAS): Validation of a Brief Version (DBAS-16). Sleep. 2007;30(11):1547–1554.

8. Emery PC. Insomnia in chronic pain patients with and without major depressive disorder. US, ProQuest Information & Learning; 2007.

9. Butcher A. Covid–19: Why sleep could be a lifesaver. The Telegraph. 7th April 2020, 2020;3rd April 2020.

10. Huang Y, Zhao N. Generalized anxiety disorder, depressive symptoms and sleep quality during COVID-19 outbreak in China: a web-based cross-sectional survey. Psychiatry Res. 2020;288:112954.

11. Casagrande M, Favieri F, Tambelli R, Forte G. The enemy who sealed the world: Effects quarantine due to the COVID-19 on sleep quality, anxiety, and psychological distress in the Italian population. Sleep medicine. 2020.

12. Harvey AG. A cognitive model of insomnia. Behav Res Ther. 2002;40:869–893.

13. Fuller PM, Gooley JJ, Saper CB. Neurobiology of the sleep-wake cycle: sleep architecture, circadian regulation, and regulatory feedback. J Biol Rhythms. 2006;21(6):482–493.

14. Potter GD, Skene DJ, Arendt J, Cade JE, Grant PJ, Hardie LJ. Circadian Rhythm and Sleep Disruption: Causes, Metabolic Consequences, and Countermeasures. Endocr Rev. 2016;37(6):584–608.

15. Altena E, Baglioni C, Espie CA, et al. Dealing with sleep problems during home confinement due to the COVID-19 outbreak: Practical recommendations from a task force of the European CBT-I Academy. J Sleep Res. 2020:e13052.

16. Czeisler CA, Duffy JF, Shanahan TL, et al. Stability, Precision, and Near-24-Hour Period of the Human Circadian Pacemaker. 1999;284(5423):2177–2181.

17. Thomas JM, Kern PA, Bush HM, et al. Circadian rhythm phase shifts caused by timed exercise vary with chronotype. JCI insight. 2020;5(3).

18. Mander BA, Winer JR, Walker MP. Sleep and Human Aging. Neuron. 2017;94(1):19–36.

19. Blackwell T, Yaffe K, Laffan A, et al. Associations of objectively and subjectively measured sleep quality with subsequent cognitive decline in older community-dwelling men: the MrOS sleep study. Sleep. 2014;37(4):655–663.

20. Peter-Derex L, Yammine P, Bastuji H, Croisile B. Sleep and Alzheimer’s disease. Sleep Medicine Reviews. 2015;19:29–38.

21. WHO. Survey tool and guidance: Behavioural Insights on COVID19. In:2020.

22. Kroenke K, Strine TW, Spitzer RL, Williams JB, Berry JT, Mokdad AH. The PHQ-8 as a measure of current depression in the general population. Journal of affective disorders. 2009;114(1-3):163–173.

23. Kroenke K, Spitzer RL. The PHQ9: A New Depression Diagnostic and Severity Measure. Psychiatric Annals. 2002;32(9):1–7.

24. Spitzer RL, Kroenke K, Williams JBW, Lowe B. A brief measure for assessing generalized anxiety disorder: the GAD-7. Archives of Internal Medicine. 2006;166(10):1092.

25. Buysse DJ, Reynolds III CF, Monk TH, Berman SR, Kupfer DJ. The Pittsburgh Sleep Quality Index: a new instrument for psychiatric practice and research. Psychiatry research. 1989;28(2):193–213.

26. Taylor MR, Agho KE, Stevens GJ, Raphael B. Factors influencing psychological distress during a disease epidemic: Data from Australia’s first outbreak of equine influenza. BMC Public Health. 2008;8(1):347.

27. Espie CA. Insomnia: Conceptual Issues in the Development, Persistence, and Treatment of Sleep Disorder in Adults. Annual Review of Psychology. 2002;53(1):215–243.

28. Reid KJ, Baron KG, Lu B, Naylor E, Wolfe L, Zee PC. Aerobic exercise improves self-reported sleep and quality of life in older adults with insomnia. Sleep medicine. 2010;11(9):934–940.

29. Morita Y, Sasai-Sakuma T, Inoue Y. Effects of acute morning and evening exercise on subjective and objective sleep quality in older individuals with insomnia. Sleep medicine. 2017;34:200–208.

30. Kudielka BM, Federenko IS, Hellhammer DH, Wüst S. Morningness and eveningness: the free cortisol rise after awakening in “early birds” and “night owls”. Biological psychology. 2006;72(2):141–146.

31. Facer-Childs ER, Middleton B, Skene DJ, Bagshaw AP. Resetting the late timing of ‘night owls’ has a positive impact on mental health and performance. Sleep medicine. 2019;60:236–247.

